# On the Kermack–McKendrick with Weibull residence times

**DOI:** 10.1101/2024.03.18.24304502

**Authors:** M. Cruz–López, A. Murillo-Salas, J.X. Velasco-Hernández

## Abstract

In this paper we develop a generalization of the Kermack-McKendrick SIR model where the time of infectiousness follows the Weibull distribution. We compute the equivalent classical results available for the classical SIR model, particularly a general expression for the basic reproduction number. We comment about the implications of this generalization in epidemic and metapopulation dynamics and illustrate our findings with some numerical simulations.

## 1 Introduction

The Kermack-Mckendrick or SIR mathematical model has been a standard for the theoretical investigation of the dynamics of directly transmitted diseases, particularly the characteristics and conditions that lead to and describe epidemic outbreaks e.g., [3, 6, 7, 12, 16]. The standard SIR model, given in Eq (1)

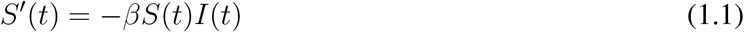

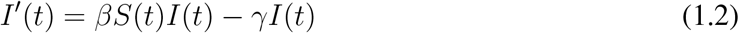

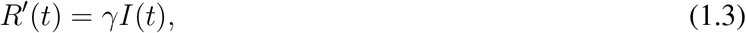

incorporates three compartments (*S* susceptible, *I* infected and *R* recovered individuals, respectively) and incorporates only two parameters: the effective contact rate *β* and the recovery rate *γ*. It is a simplification of a more general age of infection model [12] that takes into account two different time scales: the one corresponding to the chronological age of the human population and the other corresponding to the duration of infectiousness, each with their particular waiting time distributions. The simplified SIR model is framed in ordinary differential equations that, implicitly, assume that both chronological time and age of infection time have exponentially distributed waiting times. To account for the observed distribution of age of infection, the SIR framework is enlarged with one or more compartments that approximate the distribution of infectiousness in the population. There are recent results [1] that directly tackle the age of infection distribution and allow a generalization of the results of classical compartmental models.

Another important area where this kind of models has been applied is ecology. The SIR family of models (SIR, SIS, SER, SEIRS, etc.,) have a characteristic structure that makes them amenable to the study of metapopulation dynamics [8, 10]. In this context, the compartments represent patches of different classes with *S* being the empty patches, *I* the colonized and propagule producing patches, *R* the damaged or perturbed patches, and so forth. In metapopulation dynamics an important problem are the conditions and consequences of extinction events on the abundance of the species that constitutes the metapopulation.

In what follows we will use the epidemiological framework to introduce our results.

## 2 Preliminaries

The infectivity distribution is the distribution of residence times in the infected compartment. There are several ways to incorporate infectivity distributions into epidemiological models (e.g., [11, 6]). Here we follow the description presented in [14]. To motivate the construction let *x*(*τ*) be a cohort of individuals all infected at time *τ*. Individuals leave the infectious state at a rate *γ* thus

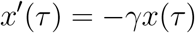

which results in

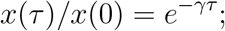

therefore, the fraction of recovered individuals at *τ* is

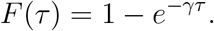

The average residence time in the infectious compartment is then

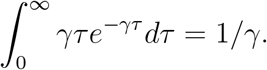

This elementary result comes from the assumption that residence times are exponentially distributed. We can provide alternative options. For example, as in [14], let *X* be an absolutely continuous random variable on [0, *∞*), of residence times with distribution *F* (*t*) = *P* [*X ≤ t*]; this leads to the survival function *G*(*t*) = 1 *− F* (*t*) = *P* [*X > t*]. Thus for *τ >* 0, *G*(*t − τ*) is the infected fraction at time *τ >* 0 that is still sick (infectious) at time *t* (*t > τ*). Armed with these observations, we can define the incidence as *βI*(*τ*)*S*(*τ*)*G*(*t − τ*) giving the new cases per unit time. The prevalence then is as

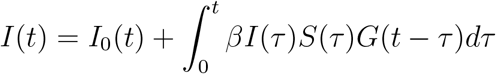

and the recovered individuals by

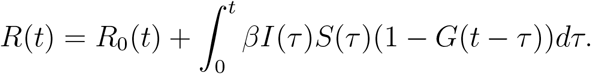

These two equations substitute the corresponding ones given in Eq. (1).

The Weibull distribution has been used to approximate incidence in epidemiological models as a good statistical descriptor of observed data [18], as a descriptor of the spreading behavior of epidemics [19] or because it provides a non-Markovian SIS model approximation on a network [15]. This last work gives results on the threshold parameter for persistence or extinction of the disease. The work [4] approximates the classical SIR model using arbitrary distributions for the infectious period, estimate the threshold parameter and fit COVID data to model simulations. All these are examples from epidemiology but the Weibull distribution can render more general dynamics that can explain processes in general ecological settings e.g., [11, 23].

In this work we are interested in characterizing the basic parameters that describe a general growth and extinction process using the Weibull distribution. We will compare them to the classical results of the SIR and other metapopulation models, particularly in relation to the shape parameter *α* of the Weibull distribution. We discuss the effect of this more realistic distribution on epidemic and ecological extinction processes.

Let us consider an absolutely continuous random variable *X* concentrated on (0, *∞*). Let *f* and *F* be its density and distribution functions, respectively. The *hazard function* associated to *X* will be denoted by *H*, and it is defined as

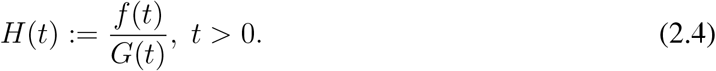

These kind of distributions can be useful in modelling random phenomena occurring in time. Note that, in case that *X* is an exponential random variable with rate *γ >* 0, we have that

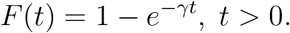

Thus, in this case the hazard function is

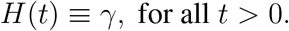

The exponential distribution is the unique continuous distribution with a constant hazard function, because this is the only continuous distribution with the lack of memory property; in other words, the *events* appear homogeneously in time.

Let *X* be a random variable with Weibull distribution, namely, with density function

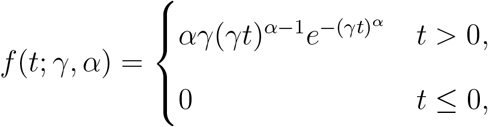

with parameters *α >* 0 (shape) and *γ >* 0 (scale). It turns out that *X* has distribution function given by

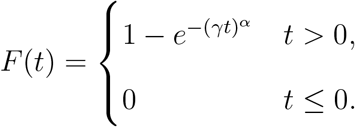

Observe that, if the shape parameter *α* = 1, *X* reduces to an exponential random variable with mean 1*/γ*. It is known that

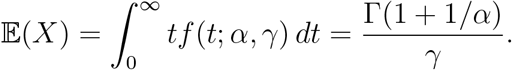

In this case, the hazard function becomes

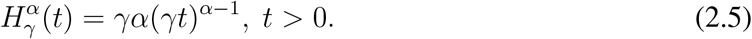

Recall that 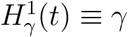, for all *t >* 0.

### Remark 1

The Weibull distribution does not have the lack of memory property but it is interesting to observe the following: depending on the value of the shape parameter *α*, the hazard function goes to 0 (0 *< α <* 1) or grows to infinity (*α >* 1), as *t → ∞*. In other words, in the first case, as time grows it is less likely to observe the occurrence of an event, whereas in the second case it is more likely to observe the occurrence of events. Thus, the parameter *α* dictates the *intensity* at which random events appears. As we noted above, the case *α* = 1 gives the exponential distribution, in which case the intensity is constant in time.

### Remark 2

In this work, an event in question is an individual ceasing to be infectious or, in a metapopulation context, an species going extinct in a given patch. In the first case *α <* 1, most individuals go extinct or abandon the infectious stage early in the process implying that most of the infections or colonization events occur early with only relatively few cases occurring in later times; in the second case *α >* 1, individuals abandon the stage later in time implying that the probability of propagation occurs at later times implying late extinction or recovery events. See Figure 2b for examples.

**Figure 1.**
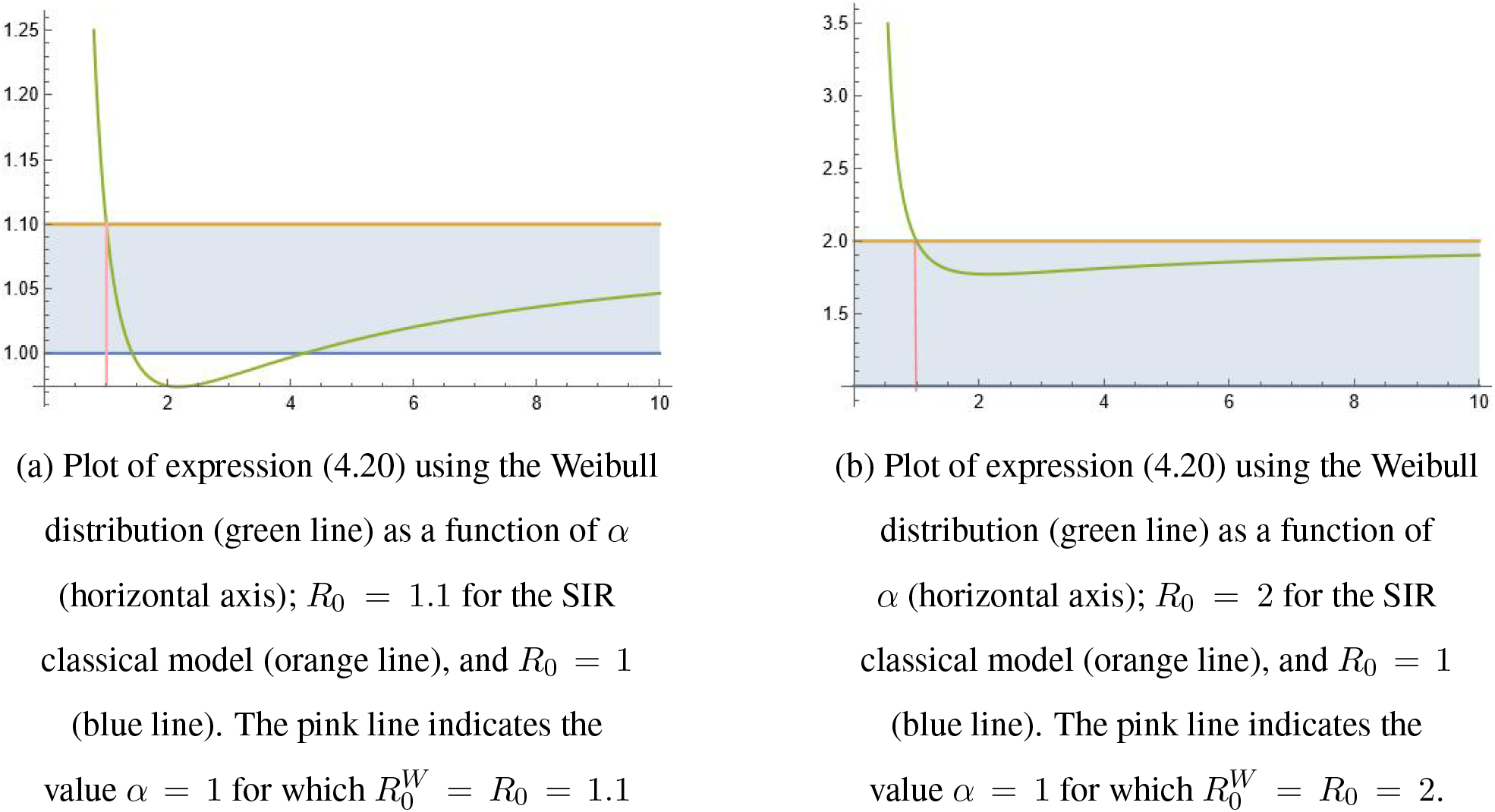
Plots of the basic reproduction number 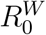

**Figure 2.**
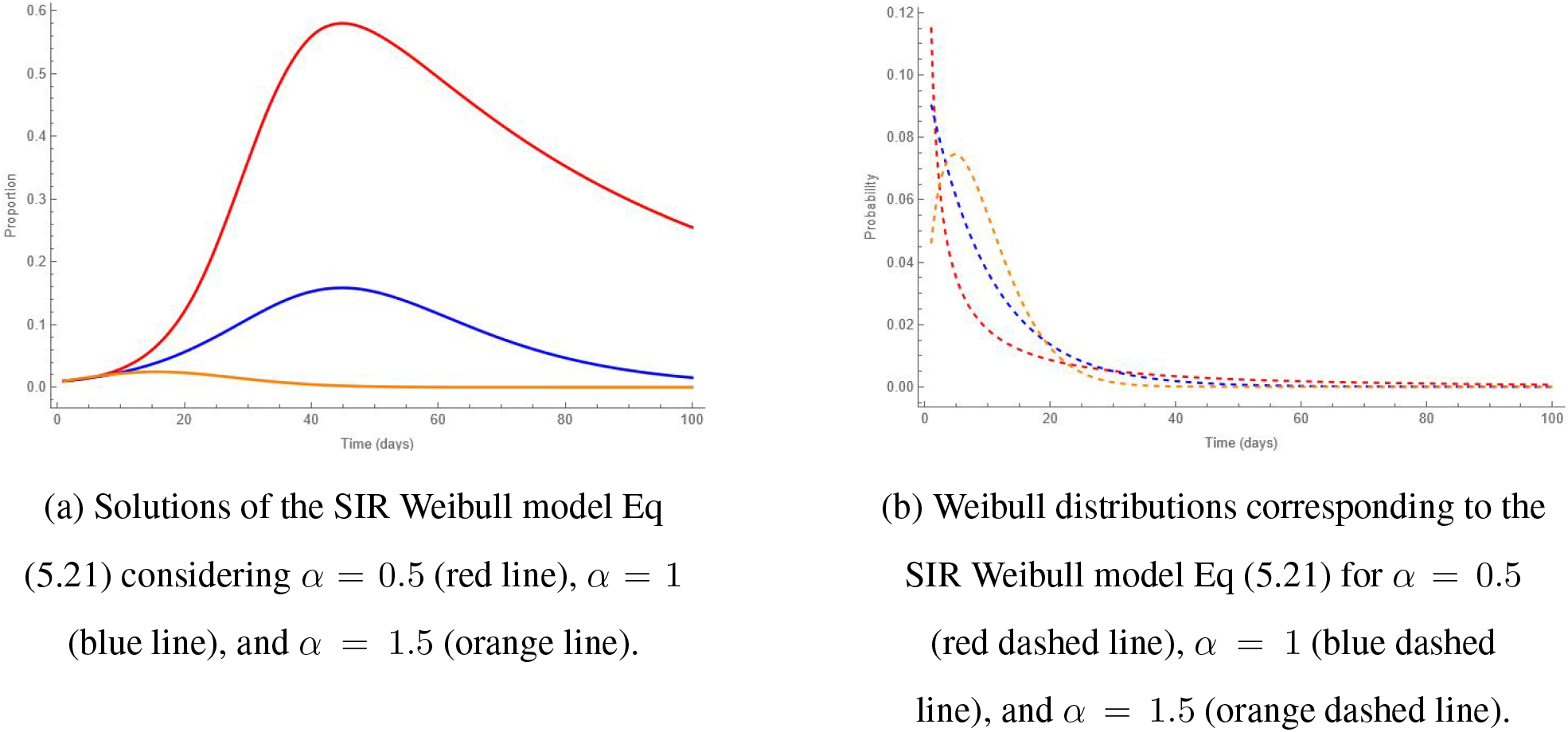
General characteristics of solutions of the SIR model with Weibull hazard rate for *R*_0_ = 2.0 (*α* = 1). The corresponding values of the reproductive number for *α* = 0.5 and *α* = 1.5 are, respectively, 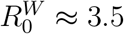, and 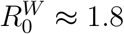

## 3 The Kermack–McKendrick model with Weibull recovering times

### 3.1 A SIS model

In this section we consider the SIS (susceptible infectious susceptible) model. As usual, the population is homogeneous, *S*(*t*) denotes the susceptible individuals at time *t, I*(*t*) are the infected individuals at time *t* and births, deaths or migrations are not considered, so *N* (*t*) = *S*(*t*) + *I*(*t*). The corresponding system of differential equations is:

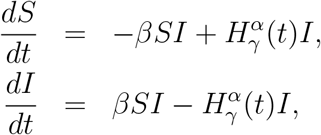

where 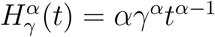. When *α* = 1 this system corresponds exactly to the classical SIS model.

Setting *N* = 1 and substituting *S*(*t*) = 1 *− I*(*t*) into the second equation we obtain

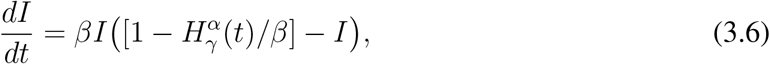

which is a logistic-type equation. Observe that the function 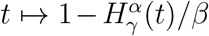 is monotone increasing or monotone decreasing depending of the value of *α <* 1 or *α >* 1, respectively. Moreover, it has a unique root at 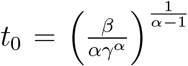. The only equilibrium points of the equation (3.6) are the points located at *I* = 0 in which case there are no infected individuals and at 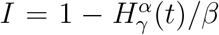. From this, we know that there exists a unique *t*^*\**^ *>* 0, with 0 *< t*^*\**^ *< t*_0_, such that the other equilibrium point is located at 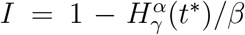. The biological relevant case occurs when 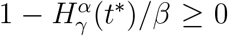. The behaviour of *I* coincides with the classical case (*α* = 1), once the value *t*^*\**^ is fixed, i.e.,

- If 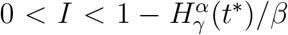, since *dI/dt >* 0, it follows that 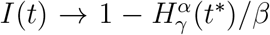 when times grows.
- If 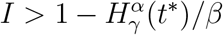, since *dI/dt <* 0, it follows that 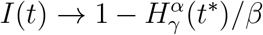 when times grows.

The susceptible population satisfies 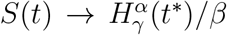 when times grows so, for all time, there will be infected and susceptible individuals rendering the disease endemic.

#### Proposition 1

*The solution to equation (3.6) is given by*

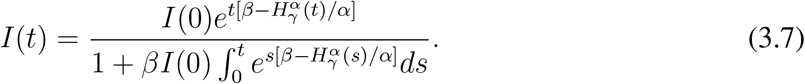

**Proof:** In order to find an explicit expression for *I*(*t*) we proceed as follows. First, we write the equation for the derivative of *I* in the form

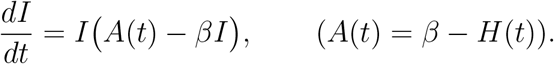

We can rewrite the equation as

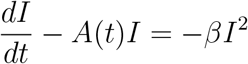

which has as general solution

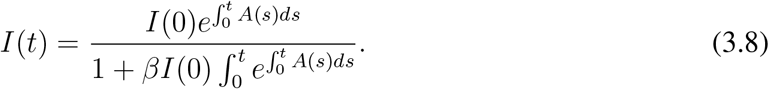

Now, since 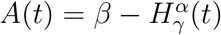 by integration we obtain:

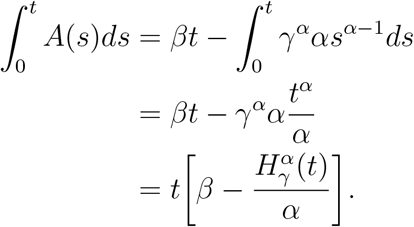

Putting this expression into the general solution (3.8) we obtain (3.7).

Note that, when *α* = 1 we recover the classical SIS solution:

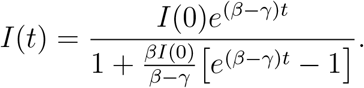

Observe that if 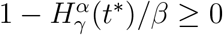 then

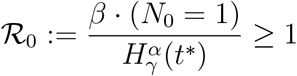

gives the *basic reproductive number*.

### 3.2 A SIR model

In this section we will use the epidemiological analogy to presents our results; later, we will make some remarks on their generalization to metapopulations. For now, let us consider a SIR model such that

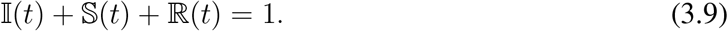

We assume that the recovering times are modeled by a Weibull random variable *τ*, i.e, with distribution function 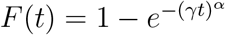, with *α, γ >* 0. We suppose that

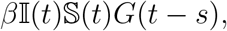

denotes the incidence, that the prevalence satisfies

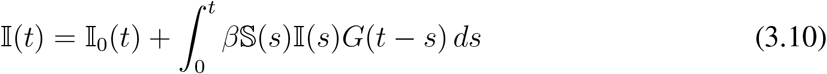

and the recovered individuals are characterized by

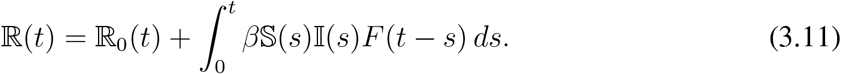

The model must satisfy the following *biological* restrictions:

- For all *t* ≥ 0, 0 ≤ 𝕀 (*t*) *≤* 1, similarly 𝕊 (*t*) and ℝ (*t*);
- _*𝕀∞*_ + 𝕊_*∞*_ + ℝ_*∞*_ = 1, 𝕊_0𝕀0_ *>* 0 and 𝕊_0_ + 𝕀_0_ = 1, since ℝ (0) = 0.

Then, from (3.9)-(3.11), is follows that the susceptible individuals satisfy

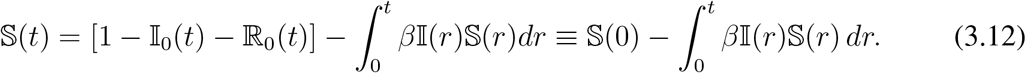

From (3.12) we have that, *t* ↦ 𝕊 (*t*) is a non increasing function. In particular, 𝕊 (*t*) ≤ 𝕊 (0) for all *t >* 0. Set 𝕊(*∞*) *≡* lim_*t*→∞_𝕊 (*t*).

Note that, taking limits in (3.12) and reordering terms, we get that

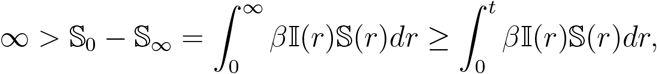

for all *t >* 0. Thus, 𝕊 (*t*) *>* 0, for all *t >* 0.

Taking derivatives in (3.12) we get that

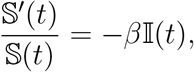

equivalently,

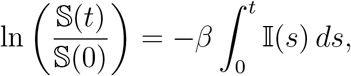

in other words,

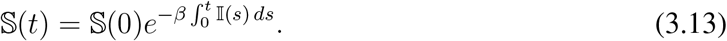

#### Theorem 1

*(Total infected population or total metapopulation size) Let X be the recovering time with distribution function F and finite mean* 𝔼 (*X*) *< ∞. Then, it holds that*

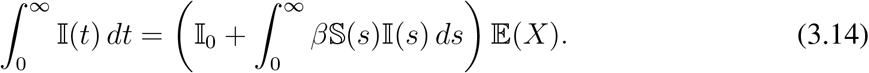

**Proof:** Note that the first term in the right-hand side of (3.10) is given by 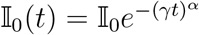. Then, integrating from 0 to *∞* we have

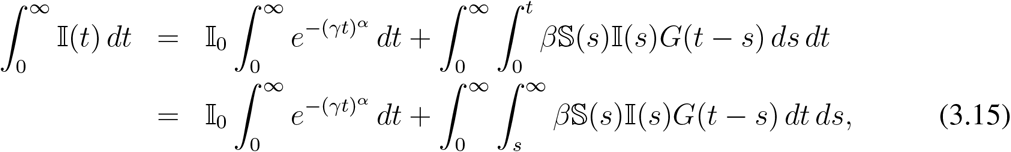

to get that last expression we have changed the order of integration. Now, observe that

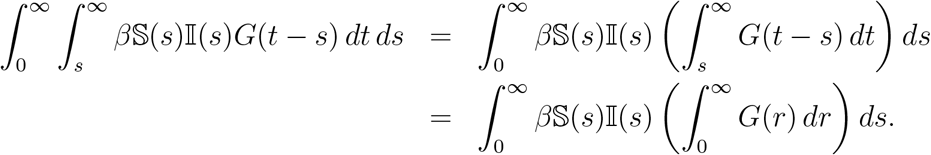

It is know that, if *X* is a random variable on (0, *∞*) with distribution function *F* then

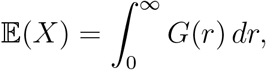

where the expectation is finite whenever the integral is finite. Hence, plugging this last expression into (3.15) we obtain (3.14).

#### Corollary 1

*Assume the conditions of Theorem 1. Then*,

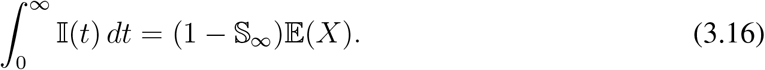

**Proof:** Taking limits equation (3.12) we can see that 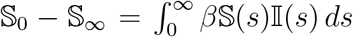. Then, using (3.14) we get that

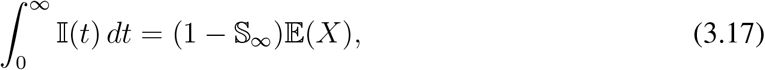

where we have the condition 𝕀_0_ + 𝕊_0_ = 1. In particular, from (3.17) we get that 𝕀 is integrable on [0, *∞*), in whose case 𝕀_*∞*_ = 0.

#### Remark 3

*It is worth to mention that Theorem 1 and Corollary 1 are valid not only for Weibull distribution but for any random variable X with finite mean*.

We recall that, in the case that *X* has a Weibull distribution with hazard function (2.5), it holds that 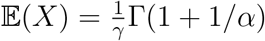. Therefore, plugin this into (3.17), we get that

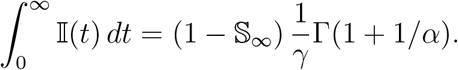

In order to compute 𝕊_*∞*_ we note that, making *t ↑ ∞* in (3.13) and using (3.17), we have that 𝕊_*∞*_ satisfies

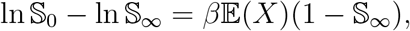

which is equivalent to

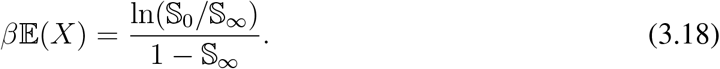

#### Remark 4

*Note that, if X has an exponential distribution with rate γ, then* 𝔼 (*X*) = 1*/γ. Therefore, from (3*.*18) we have that* 𝕊_*∞*_ *satisfies*

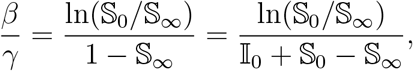

*where we have used the relation* 1 = 𝕀_0_ + 𝕊_0_, *which coincides with the classical case*.

## 4 *R*_0_ for a general waiting time distribution

In this section we compute the basic reproduction number which, in a general framework, represents the propagule and colonization potential during the time that a colonized patch has not gone extinct. By (21) from [11]

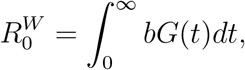

where 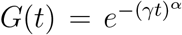 denotes the probability that a newly infected individual (newly colonized patch) remains infected (occupied) by time *t* and *b ≡ βS*(0) is the average of new infected individuals (new colonized patches) infected (colonized) by an infected (colonized) one. Therefore,

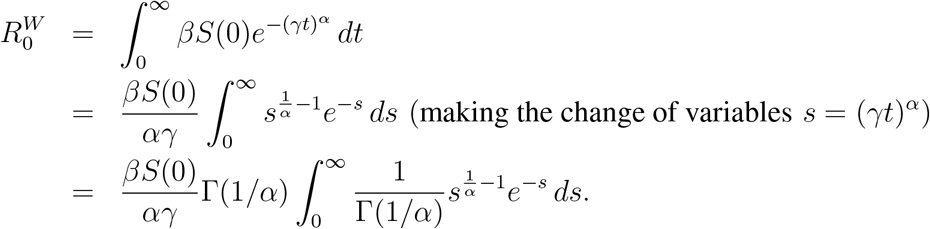

Finally, we conclude that

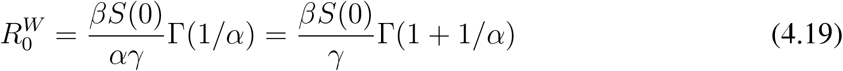

Note that, for *α* = 1, we recover that

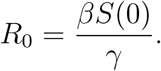

### Remark 5

Note that 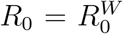 when *α* = 1 and that 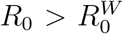 for *α >* 1 and 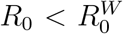 when*α <* 1. So we have two interesting possibilities. If *R*_0_ *<* 1 then 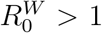 for *α <* 1 implying the existence of epidemic outbreaks (patch colonization) in the Weibull case when the classical SIR model predicts no outbreak (no successful colonization of patches). The other case occurs when *R*_0_ *>* 1 but it is relatively low. In this case it can be that 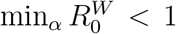 implying no epidemic outbreak (unsuccessful colonization) in the Weibull case. In summary for *R*_0_ *>* 1 but low, there exists an interval 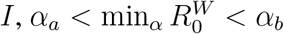, such that if *α ∈ I* then 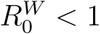.

### Remark 6

Observe that the computations that leads to (4.19) can be done for a general random variable *X* with distribution function *F*. To be more precise, if we denote by 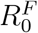 the reproduction number with distribution function *F*, we obtain that

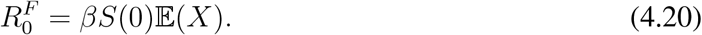

Thus, if *X* has an exponential distribution with mean 1*/γ* we get that 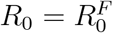.

We conclude this section pointing out the biological significance of expression (4.20) for it gives a very general, and relatively simple, formula for the reproductive number. Figure 4a shows 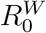 as a function of the parameter *α*. As it can be seen, 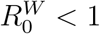 in the region between the green curve and the horizontal blue line, but 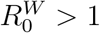 the rest of the interval. In the Figure, 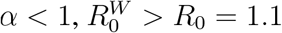 and for 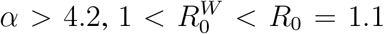. Figure 4b shows a different behavior. Here *R*_0_ = 2 (in the SIR classical model) and then 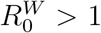 always. In fact, the minimum value of *R*_0_ that renders 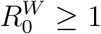 for all *α >* 1 is *R*_0_ *≈* 1.13. Therefore, in this case 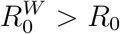 for *α <* 1 and for *α >* 1, it is always greater but less than *R*_0_ = 2.

We have then that, if in an epidemic (colonization) event the large majority of cases is concentrated during the first days of the process, the associated reproductive number can be significatively larger than the one predicted by the classical *R*_0_ of the SIR model; however, if the transmission (propagation) events occur mostly at later times in the process, the associated reproductive number will be smaller than the *R*_0_ of the classical SIR model. So, the higher the SIR *R*_0_ reproductive number, the less difference with 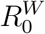 for large *α*.

## 5 Conclusions

As a manner of conclusion on the approach that we have taken in this work, we now look at a simple and straightforward incorporation of non-exponentially distributed waiting times to the SIR model. We want to keep the nature of the model system, ordinary differential equations, but introduce a distribution of waiting times, that is of infectivity times, different from the exponential.

To this end let *β* and *γ* be the effective contact and cure rates, respectively, of the classic SIR model. As we have seen in the previous sections, 1*/γ* is the expected value of the (constant) exponential waiting time of individuals in the infectious compartment, in other words, 1*/γ* is the mean infectivity period. This period is constant because we are assuming exponentially distributed waiting times. We change this assumption in the follwing model

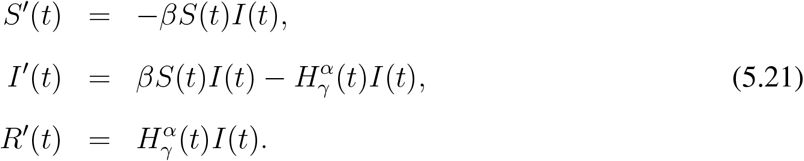

Thus, we can think of this system as the *Kermack–McKendrick model with infectivity times modeled by a Weibull distribution* with hazard rate given by 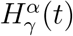. When *α* = 1, we recover the classical SIR model. The system (5.21) is a non-autonomous differential equation that, as the classical model, still has as equilibria the points *E*(*S, I*) = (*S*^*\**^, 0) for *S*^*\**^ *∈* [0, 1] and thus can be easily comparable. The reproductive number can be computed by substituting the expected (constant) waiting time 1*/γ* from the exponential distribution, by the expected waiting time of the Weibull distribution, rendering

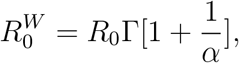

where *R*_0_ = *βS*(0)*/γ*.

Figure 2 shows the behavior of three different epidemics for different values of *α* with *R*_0_ = 2 (corresponding to the case *α* = 1). It can be seen that 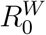 changes slowly if *α* is increased slightly (from 1 to 1.5 in this example) but a reduction in the same amount can have a dramatic impact on the value of 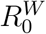 and, therefore, in the size of the epidemic. As it can be seen from the figure, the time to the maximum peak size does not vary much for *α* = 1 and *α* = 0.5 but the relative size of the outbreak does change a lot (from 16% to 58% prevalence).

If we look a little closer at Figure 2 for the case *α >* 1 (see Figure 3), we can appreciate that the initial increase in prevalence is slower and nearly linear compared to the decline in prevalence after the peak which occurs in an exponential way. Slow, almost linear growth was observed during the COVID-19 pandemic [22]. An explanation, presented in the previos cited work is that this growth pattern can be understood as a consequence of the structure of low-degree contact networks resulting from the extended use of non-pharmaceutical interventions (NPI). The Weibull approach presented here qualitatively agrees with this explanation if we assume that the net effect of NPIs is to delay the bulk of infectious processes later in time, forcing the parameter *α* to be larger than one, i.e., essentially changing the exponential distribution of waiting times to a Weibull distribution.

**Figure 3.**
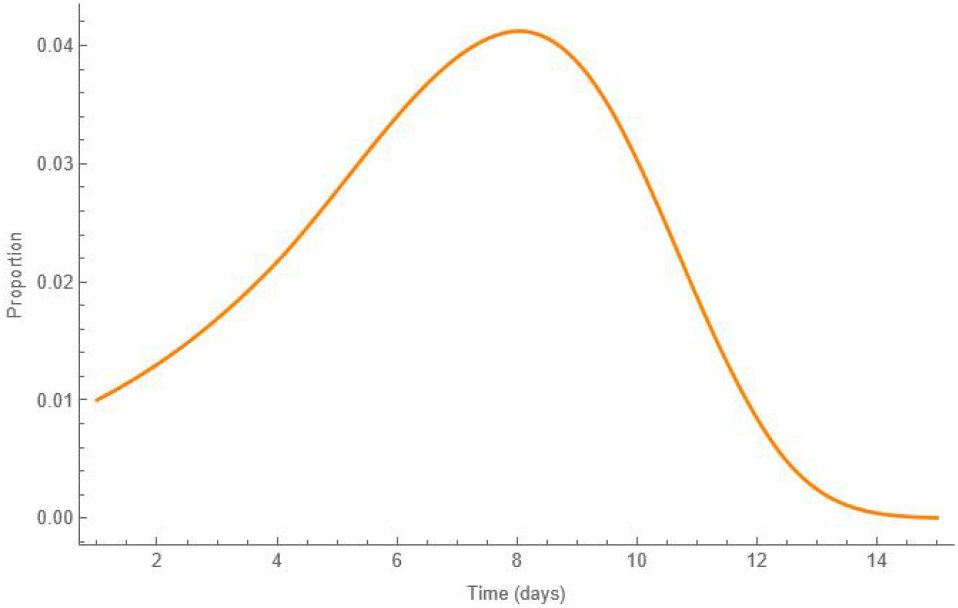
Numerical solution of the SIR with Weibull hazard function for *α* = 5

Moreover, the COVID-19 pandemic illustrated the role of superspreading events on the development and fate of the epidemic [2, 5, 13, 17, 20] which together with the action of non-pharmaceutical interventions and other mitigation measures affected the speed of contagion along the evolution of the epidemic. This changes in speed can be described statistically [21, 9, 19]. Looking at these changes in transmission speed as extreme transmission events, then their theoretical distribution can be described by either of the standard distributions that are used to model extreme events (e.g., [9]). In this paper we present some results on the properties of a SIR equations system that uses the Weibull distribution to model the waiting times of an individual in the infectious stage. Our main results provide a general formula for the reproductive number that can be generalized to other distributions of waiting times.

## Data Availability

All data produced in the present work are contained in the manuscript

## Acknowledgments

JXVH acknowledges support from UNAM PAPIIT grant IV100220.

